# Development and Validation of a Deep Survival Model to Predict Time-to-Seizure from Routine EEG

**DOI:** 10.1101/2025.08.12.25333063

**Authors:** Émile Lemoine, An Qi Xu, Mezen Jemel, Frédéric Lesage, Dang K. Nguyen, Elie Bou Assi

## Abstract

**Objective:** To develop and validate a deep survival model (EEGSurvNet) that analyzes routine EEG to predict individual seizure risk over time, comparing its performance to traditional clinical predictors such as interictal epileptiform discharges (IEDs).

**Methods:** We conducted a retrospective cohort study including 1,014 consecutive routine EEGs from 994 patients recorded at a tertiary epilepsy center. We developed EEGSurvNet, a deep learning model that predicts time-to-next-seizure over a two-year horizon from a single EEG. Model performance was evaluated on a temporally-shifted testing set of 135 EEGs from 115 patients using time-dependent area under the receiver operating characteristic curve (AUROC), AUROC integrated over two years (iAUROC), and C-index. We compared the deep survival model to a clinical Cox model incorporating standard risk factors as well as a random model based on baseline seizure risk.

**Results:** EEGSurvNet achieved a two-year iAUROC of 0.69 (95%CI: 0.64–0.73) and C-index of 0.66 (0.60–0.73), outperforming both clinical and random models. Performance was highest in the first months following EEG, peaking at 2 months (AUROC = 0.80). Combining EEGSurvNet to clinical predictors further improved performances (iAUROC = 0.70, C = 0.69). Notably, the model showed superior discrimination on EEGs without IEDs (iAUROC = 0.78 vs 0.53). Model interpretation revealed that the temporal-occipital regions and 6–15 Hz frequencies contributed most to risk prediction.

**Significance:** EEGSurvNet demonstrates that deep learning can extract prognostic information from routine EEG beyond visible epileptiform abnormalities, potentially improving patient counseling and treatment decisions. Future prospective studies are needed to validate these findings and assess their clinical impact.

**Key Points:**

- Deep learning model predicts individual seizure risk from routine EEG over 2 years
- Model performs better on EEGs without epileptiform discharges, suggesting novel biomarkers
- Temporal-occipital regions and 6-15 Hz frequencies contribute most to risk prediction
- Combined clinical-EEG model achieves best performance for seizure risk stratification

## Introduction

Epilepsy is practically defined by an increased risk of seizures.^1^ Risk assessment requires a multimodal approach integrating the semiology of suspicious episodes, clinical risk factors, neuroimaging, and electroencephalogram (EEG). The presence of interictal epileptiform discharges (IEDs) on EEG is particularly informative, predicting a 1.5–3 times higher recurrence risk in various clinical situations.^2–5^

Despite its central role in epilepsy, EEG has several limitations. Its sensitivity for epileptiform anomalies is low, with only 29–55% of patients with epilepsy showing IEDs on a 30–60-minute EEG.^5–7^ Its interpretation requires subspecialized expertise, and even among experts, inter-rater agreement for epileptiform abnormalities is at best moderate.^8,9^ Computational EEG analysis, particularly deep learning, offers a promising alternative by automatically extracting features that represent complex interactions between frequencies and brain regions across different time scales.^10–13^

Current automated EEG analysis models primarily focus on diagnostic classification ^10,11,14,15^ or predicting recurrence at fixed time points.^16,17^ This approach does not fully reflect the temporal nature of epilepsy risk.^1,18^ Survival analysis estimates the survival function *S*(*t*) describing disease evolution after exposure to a risk factor or treatment.^19^ Survival models are particularly suited for epilepsy to model seizure risk over time, but they are typically limited by the low complexity of their input, often consisting of few clinical variables.^20–22^ However, when coupled with deep learning, survival models can map complex input to time-dependent risk prediction.^23,24^ Several studies have demonstrated the utility of deep survival models in oncology^24–26^ and intensive care,^27^ but their applicability in epilepsy remains unexplored.

This study proposes a deep survival model in epilepsy, EEGSurvNet, which analyzes the EEG signal to predict time to next seizure over a two-year horizon. The model’s performance is compared to a traditional survival model with standard clinical predictors, including the presence of IEDs. Beyond prediction, we explore model interpretability to identify EEG signal characteristics associated with seizure risk and parameters that optimize its generalization.

## Methods

### Study Design and Population

This is a retrospective study of consecutive patients who underwent routine EEG at the Centre hospitalier de l’Université de Montréal (CHUM), Canada. All patients who had an EEG in the clinical neurophysiology unit between January 1, 2018, and December 31, 2019, were included. Routine EEG includes 30–60-minute recordings with and without sleep deprivation performed in outpatient settings. Exclusion criteria were absence of follow-up after EEG, uncertain epilepsy diagnosis at the end of follow-up, or seizures during EEG recording. Patient records were reviewed by a neurology resident (EL) and three neuroscience students (MJ, AQX, JDT) following a pre-specified protocol. Extracted data included age, sex, comorbidities, epilepsy risk factors, and neuroimaging findings. For each visit, the number of seizures since the previous visit was extracted. Epilepsy diagnosis was determined according to the International League Against Epilepsy criteria^1^ and relying on the last available treating neurologist’s note. EEG reports were reviewed for any mention of IEDs or abnormal slowing. Clinical data were stored in a REDCap database.

EEGs recorded before September 2019 constituted the training and validation sample, while those after September 2019 formed the test sample (**Figure 1A**). Patients who had EEGs both before and after September 2019 were excluded from the test sample.

**Figure 1:**
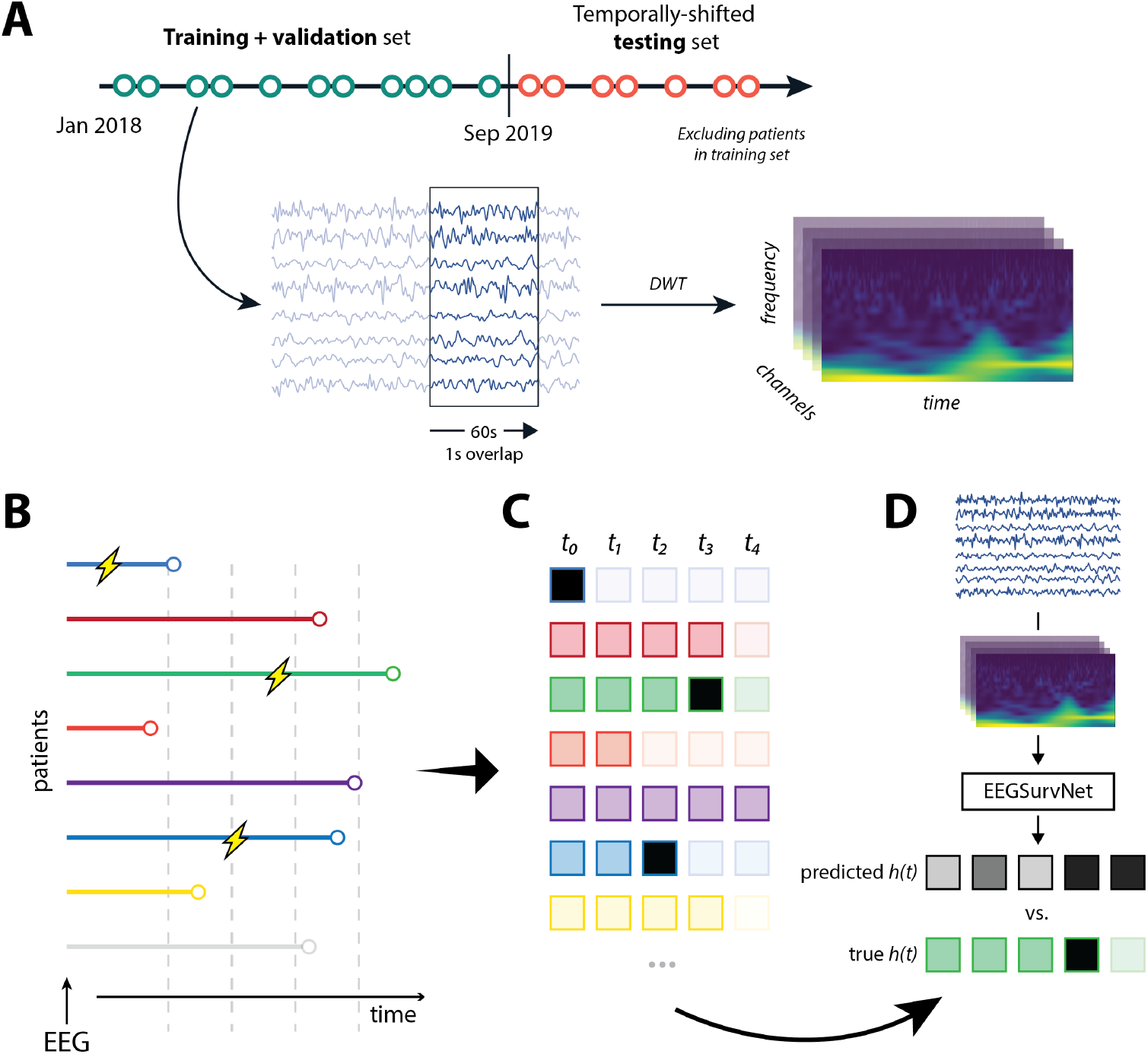
EEG processing and discrete-time survival analysis. **A**: The cohort is split into a training/validation and testing sets, without patient overlap. EEGs are segmented into 60s windows (1s overlap) and transformed into spectrograms with discrete wavelet transform (DWT). **B**: Original time series for each patient, some experiencing a seizure during follow-up (yellow lightning bolt). **C**: Temporal discretization of each patient’s follow-up, where each time-period is represented by a binary value (0: no seizure during period; 1: seizure during period). **D**: The EEGSurvNet model takes EEG signals transformed into spectrograms as input, then predicts a risk value ***h***_***t***_ for each period. Optimization uses the negative log-likelihood of the hazard model. Predictions are subsequently aggregated across segments and periods to obtain a global risk score.

### Primary outcome

The primary outcome was time (in days) to next epileptic seizure after EEG, extracted from follow-up medical notes. The outcome encompassed all types of epileptic seizures, including focal seizures with preserved consciousness and myoclonic seizures, but not non-epileptic events. For approximately 25% of patients, the exact date was not reported; in these cases, we performed linear interpolation based on the reported seizure frequency, assuming a uniform distribution of seizures over the period. Patients who did not experience seizures during their follow-up were considered censored at the date of their last documented visit. The outcome for the testing cohort was only accessed during validation.

### Predictors

#### EEG

EEGs were recorded on a Nihon-Kohden system following Canadian guidelines.^28^ Awake EEGs, lasting 20-30 minutes, were recorded at 200 Hz using 19 electrodes in the 10-20 system. They included two 90-second periods of hyperventilation (except in patients over the age of 80, non-cooperative patients, or those with medical contraindications) and photic stimulation from 4 to 22 Hz. For sleep EEGs, patients were instructed to sleep no more than half of their habitual sleep duration the night prior. Sleep EEGs lasted 60 minutes and included the same activation procedures. EEGs were recorded with a linked-ear system reference, converted to EDF format, and stored following the BIDS standard.^29^

#### Deep Survival Model

For this study, we developed a deep survival model for EEG called EEGSurvNet. EEGSurvNet is built upon DeepEpilepsy, a Vision Transformer (ViT) that takes multi-channel EEG segments of 10 or 30s as input^11^. Several improvements were made to the original architecture to enhance its performance, robustness to artifacts, and interpretability. These improvements were iteratively tested on both the training/validation set and the Temple University Abnormal EEG corpus (v3.0.1).^30^ The model architecture is described in **Supplementary Methods 1 and Supplementary Table 1**.

The model’s adaptation to survival analysis uses a discrete-time analysis approach (**Figure 1B–D**).^31,32^ The follow-up duration is divided into seven logarithmically spaced periods between 28 days and 2 years (the last period corresponding to > 2 years). The model predicts the hazard (instantaneous risk) *h*(*t*) of seizure for each period via a logistic function applied to the network outputs (Φ):

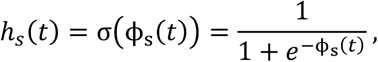

where *s* represents the analyzed EEG segment.

For training, each patient is represented by two values: the index *t*_*i*_ of the period where a seizure occurs or the last available follow-up period, and an event indicator δ distinguishing seizures (1) from censoring (0). The loss function corresponds to the negative log-likelihood of the hazard model.

Training is performed on overlapping 60-second segments (1s overlap), totaling approximately 1,800 segments per 30-minute EEG. The model is trained on two A6000 GPUs for 25 epochs, each epoch containing about 1.7M segments. Data augmentation is applied using TrivialAugment^33^ with 90% probability, adding either random Gaussian noise, random masking, or random equalization. Optimization uses cosine decay of the learning rate after a one-epoch warmup period. Hyperparameters are optimized on the validation set (**Supplementary Table 2)**. PyTorch library (v2.6.0) is used to train the deep models.

For inference on a complete EEG, we perform two levels of aggregation. First, hazards are averaged across all segments. Then, to regularize the output, we calculate a constant hazard *h*^∗^ by averaging hazards over all periods. From this constant hazard, we calculate the survival function:

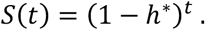

The global risk score *R* equals the constant hazard *h*^∗^ and allows comparison with proportional hazard models like Cox.

### Cox Proportional Hazards Model Using Clinical Predictors

For comparison, we used a Cox proportional hazards model using traditional seizure risk predictors: age, sex, family history of epilepsy, focal lesion on neuroimaging, and IEDs on EEG. Based on previous work,^5^ we included interaction terms between the presence of IEDs and other predictors. The Cox model predicts a proportional hazard according to the equation:

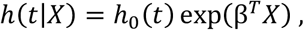

where *h*_0_(*t*) is the baseline hazard, *X* is the predictor vector, and β are the estimated coefficients. The total risk is calculated by combining the baseline risk (estimated on the training sample) with the proportional risk, producing a measure analogous to the deep model’s global risk score. The Cox model was fitted on the training/validation cohort.

### Combination of Clinical and Deep Models

A third approach combines predictions from both deep and clinical models by multiplying their respective risk scores:

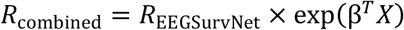

### Sample Size and Power Analysis

Power analysis was performed using the R library “powerSurvEpi” following Schmoor et al.’s method.^34^ We estimated the required sample size for a Cox model with a binary predictor, representing the presence or absence of EEG abnormality. The analysis assumes an event rate (seizures) of 0.5, a hazard ratio of 2.0, a significance level α of 0.05, and a target power of 0.8. According to these parameters, a sample of 131 patients is necessary to detect the effect of interest.

### Analysis

Model evaluation assessed discrimination and calibration. Discrimination was evaluated using cumulative/dynamic time-dependent ROC curve.^35^ Sensitivity and specificity are time-dependent; cumulative cases include all individuals who experienced an event up to time *t*, while dynamic controls are those who experience an event after *t*. The cumulative/dynamic AUC quantifies the model’s ability to distinguish subjects who will have a seizure before a given time from those who will have one after. We also applied inverse probability of censoring weighting (IPCW) to handle censoring bias (based on the outcome distribution in the training/validation set).^36^ We calculate AUROC at each period *t* plus the integrated AUROC over two years (iAUROC). For calibration, we used the integrated Brier score over two years.^37^ 95% confidence intervals (CI) are calculated using bootstrap (1,000 iterations).

In addition, models were compared to a random reference model centered around the baseline seizure recurrence probability observed in the training set using the Brier Skill Score (BSS). The BSS ranges from −∞ to 1, where positive values denote performance that exceeds the reference (**Supplementary Methods 2**).

To align with previous literature on non-DL seizure recurrence prediction models,^20–22,38–40^ we also calculated iAUROC and iBS at 1 year, as well as Harrell’s C-index.^41^

Stratified analyses were performed to evaluate performance in clinically relevant subpopulations: age group (18-40, 40-60, and >60 years), sex, focal lesion on imaging, abnormal EEG slowing, and IEDs.

Model interpretation and ablation analyses were performed. For interpretation, we used gradient-based Shapley values to identify EEG features contributing to predictions. The ablation study systematically evaluated key model parameters including segment duration, time and frequency resolution, and data augmentation. Details are presented in **Supplementary Methods 3**.

### Standard Protocol Approvals, Registrations, and Patient Consents

This study received approval from the Research Ethics Board of the Centre de recherche du CHUM (Montreal, Canada, project number: 19.334). The committee granted a waiver of informed consent due to the absence of diagnostic/therapeutic intervention and minimal risk to participants. All methods comply with Canada’s Tri-Council Policy Statement on the Ethical Conduct for Research Involving Humans. We confirm that we have read the Journal’s position on issues involved in ethical publication and affirm that this report is consistent with those guidelines.

The study source code will be available upon publication at: **https://gitlab.com/chum-epilepsy/epi_surv**. Anonymized data will be made available to qualified investigators upon reasonable request, conditional to the approval by our REB. The TRIPOD checklist is provided as Supplementary material.

## Results

### Participants

Of 1,540 EEGs performed at CHUM (1,286 patients), 1,014 EEGs from 994 patients were included: 879 EEGs from 786 patients in the training sample and 135 EEGs from 115 patients in the test sample. Median follow-up was 2.2 years after EEG for both cohorts. Clinical characteristics are presented in **Table 1**.

**Table 1:** Patient characteristics in the training and testing cohorts.

|  | Training/validation cohort | Testing cohort |
| --- | --- | --- |
| Number of EEGs (number of patients) | 879 (786) | 135 (115) |
| Seizure recurrence during follow-up (%) | 295 (33.6) | 40 (29.6) |
| Sex = woman (%) | 450 (51.3) | 73 (54.1) |
| Age (median [IQR]) | 49.00 [32.00, 62.00] | 51.00 [30.00, 63.50] |
| Total follow-up after EEG in weeks (median [IQR]) | 116.00 [52.00, 153.00] | 113.00 [57.00, 194.50] |
| Epilepsy type (%) |  |  |
| Focal | 409 (46.5) | 58 (43.0) |
| Generalized | 103 (11.7) | 13 (9.6) |
| Unknown | 53 (6.0) | 3 (2.2) |
| Pas d'épilepsie | 314 (35.7) | 61 (45.2) |
| History of febrile seizure (%) | 21 (2.4) | 6 (4.4) |
| Family history of epilepsy (%) | 63 (7.2) | 15 (11.1) |
| Days since last seizure (median [IQR]) | 225.00 [52.00, 1130.00] | 200.00 [73.00, 938.25] |
| Number of ASM (%) |  |  |
| 0 | 327 (37.2) | 67 (49.6) |
| 1 | 338 (38.5) | 37 (27.4) |
| 2 | 145 (16.5) | 18 (13.3) |
| 3 | 54 (6.1) | 8 (5.9) |
| 4 | 12 (1.4) | 5 (3.7) |
| 5 | 3 (0.3) | 0 (0.0) |
| Focal lesion on brain imaging (%) | 332 (37.8) | 52 (38.5) |
| Sleep deprived EEG (%) | 126 (14.3) | 20 (14.8) |
| IED (%) |  |  |
| Absence | 654 (74.4) | 103 (76.3) |
| Presence | 156 (17.7) | 26 (19.3) |
| Uncertain | 69 (7.8) | 6 (4.4) |
| Abnormal slowing on EEG (%) | 266 (30.3) | 42 (31.1) |

Seizures occurred after 295 of the training EEGs (33.6%) and 40 of the test EEGs (29.6%), with a one-year seizure-free survival rate of 69% (95%CI: 66–73%) and 72% (66–79%), respectively. Clinical predictors were similarly distributed between the training and testing cohorts (**Table 1**).

### Model Development and Training

The training/validation cohort was randomly split into a training sample (80%, 1.4M segments) and a validation sample (20%, 0.3M segments). The validation set was used to optimize the following hyperparameters: batch size, learning rate, weight decay, dropout, gradient clipping, and augmentation probability. The TUH Abnormal Corpus dataset (3.7M segments) was used to optimize the model’s input format and correct bugs.^30^ The final model was trained on the entire training/validation cohort for 25 epochs.

### Model Performance

An example of EEGSurvNet model predictions is illustrated in **Figure 2**, and complete results are presented in Table 2. On the test set, EEGSurvNet achieved a two-year iAUROC of 0.69 (95% confidence interval: 0.64-0.73) and a C-index of 0.66 (0.60-0.73). In comparison, the Cox model’s iAUROC was 0.61 (0.56-0.65; C = 0.61 [0.55-0.68]), and the combined model’s iAUROC was 0.70 (0.66-0.74; C = 0.69 [0.65-0.73]). For all these models, performance was superior to the random model (iAUROC = 0.54 [0.49-0.59], C = 0.53 [0.45-0.61]).

**Table 2:**
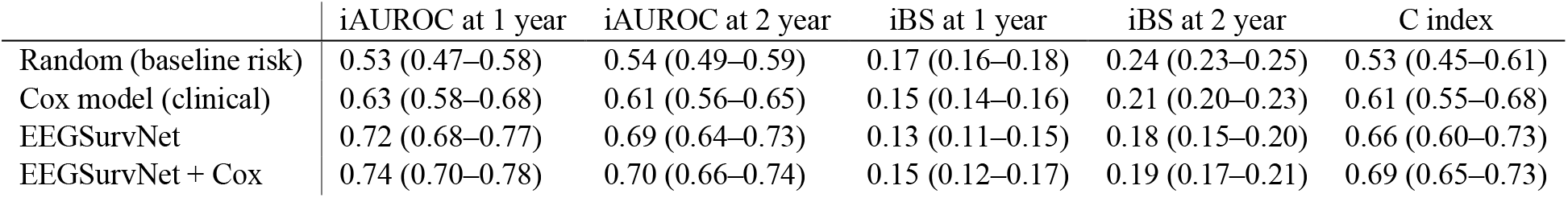
Models performance for predicting seizure recurrence risk after EEG.

|  | iAUROC at 1 year | iAUROC at 2 year | iBS at 1 year | iBS at 2 year | C index |
| --- | --- | --- | --- | --- | --- |
| Random (baseline risk) | 0.53 (0.47–0.58) | 0.54 (0.49–0.59) | 0.17 (0.16–0.18) | 0.24 (0.23–0.25) | 0.53 (0.45–0.61) |
| Cox model (clinical) | 0.63 (0.58–0.68) | 0.61 (0.56–0.65) | 0.15 (0.14–0.16) | 0.21 (0.20–0.23) | 0.61 (0.55–0.68) |
| EEGSurvNet | 0.72 (0.68–0.77) | 0.69 (0.64–0.73) | 0.13 (0.11–0.15) | 0.18 (0.15–0.20) | 0.66 (0.60–0.73) |
| EEGSurvNet + Cox | 0.74 (0.70–0.78) | 0.70 (0.66–0.74) | 0.15 (0.12–0.17) | 0.19 (0.17–0.21) | 0.69 (0.65–0.73) |

**Figure 2:**
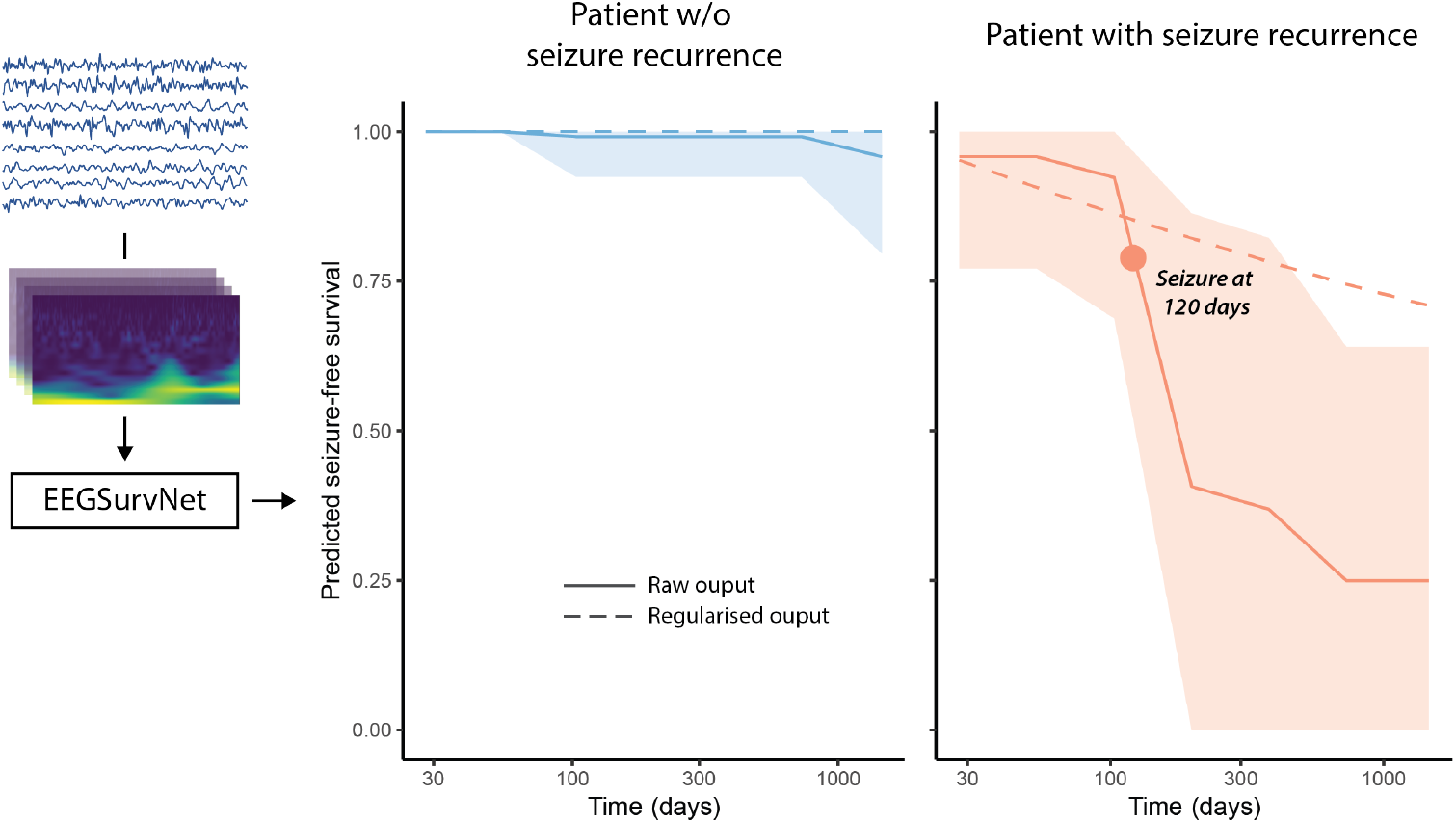
Illustration of raw predictions from the EEGSurvNet model for two patients from the test cohort. EEGs transformed into spectrograms serve as model input. The model output is then converted into a survival function that characterizes seizure-free survival at each period (***S***(_***t***_)). The raw prediction is based on hazard averaged across segments 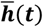, and the regularized prediction on constant risk ***h***^∗^. The shaded ribbon shows the standard deviation of predictions across segments from the same EEG.

Time-dependent AUROC is presented in **Figure 3**. For EEGSurvNet, these peak at ∼2 months (AUROC: 0.80 [0.72-0.88]) and is statistically superior to the random model between the 3^rd^ and 6^th^ months.

**Figure 3:**
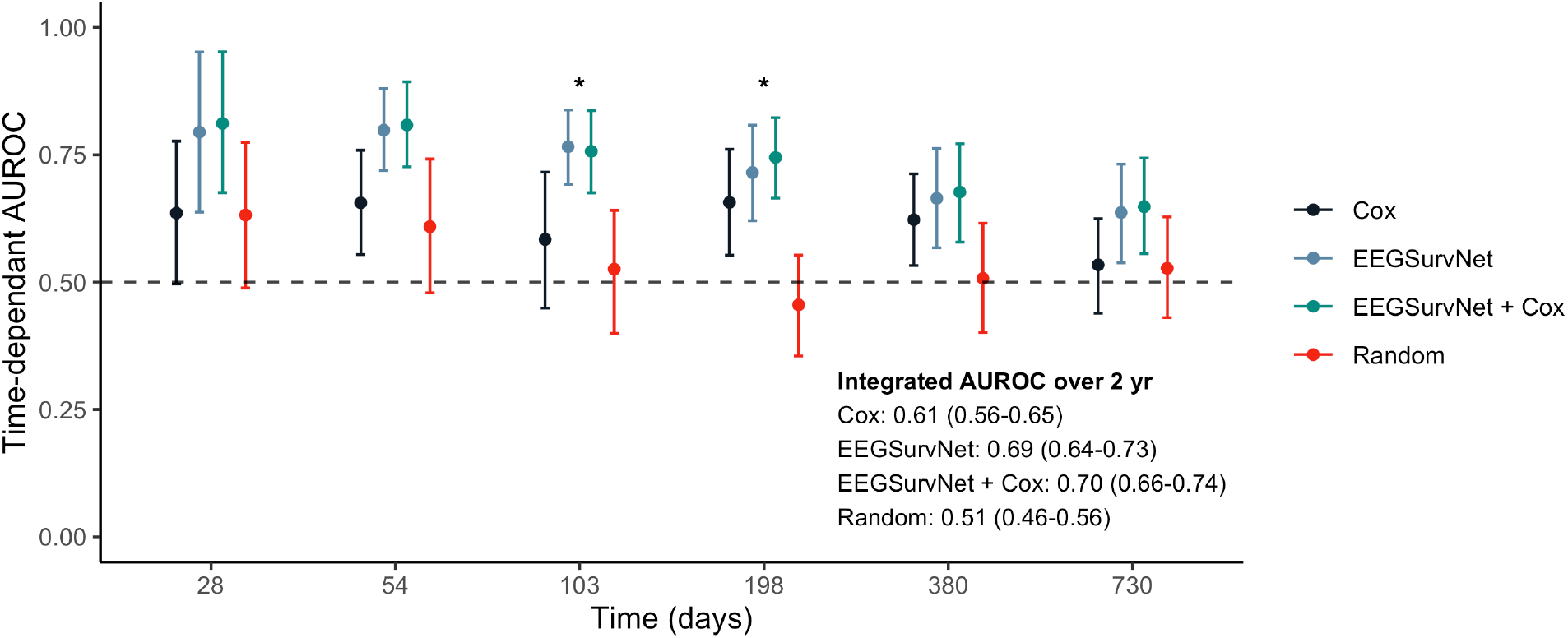
Performance of different models in predicting seizure-free survival over time. The random model corresponds to predictions based on the baseline risk of seizure recurrence drawn from the training cohort. AUROC: Area Under the ROC Curve.

For calibration, EEGSurvNet achieved an integrated Brier score at 2 years of 0.18 (0.15-0.20), significantly superior to the random model (0.24 [0.23-0.25]) (Table 2). The Brier Skill Score, measuring performance gain against the random model, is positive across all periods and peaks at 3 months with a value of 0.22 (**Figure 4**).

**Figure 4:**
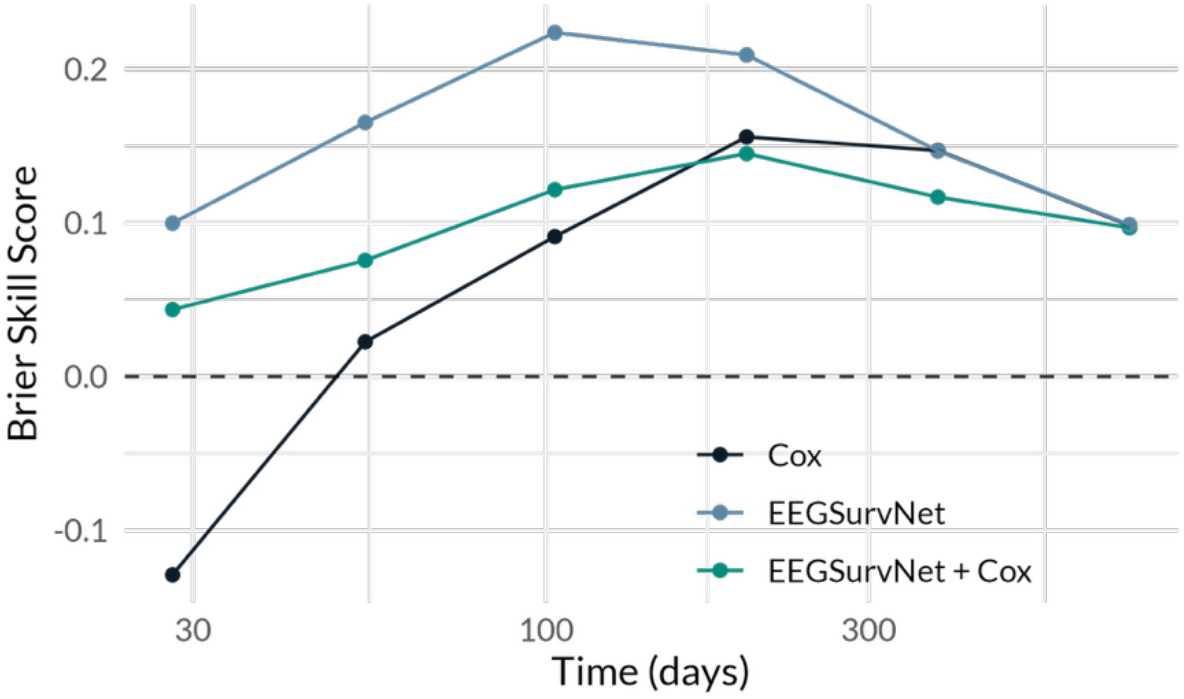
Model calibration through time. The Brier Skill Score is a measure of calibration improvement against the random model, where values between 0 and 1 indicate a calibration gain

To regularize the deep model’s output, the time-dependent risk 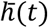 is averaged across periods for a constant risk *h*^∗^. Without this operation, EEGSurvNet’s performance is as follows: two-year iAUROC of 0.60 (0.55-0.65), a two-year iBS of 0.18 (0.16-0.20), and C-index of 0.61 (0.53-0.69).

By stratifying the test cohort according to the predicted mean survival score, we obtain three risk groups: “Low risk,” “Medium risk,” and “High risk.” Seizure-free survival of these patients is significantly associated with the risk predicted by EEGSurvNet (p = 0.003, **Figure 5**). In low-risk patients, the two-year recurrence rate is 12%, compared to 42% for high-risk patients.

**Figure 5:**
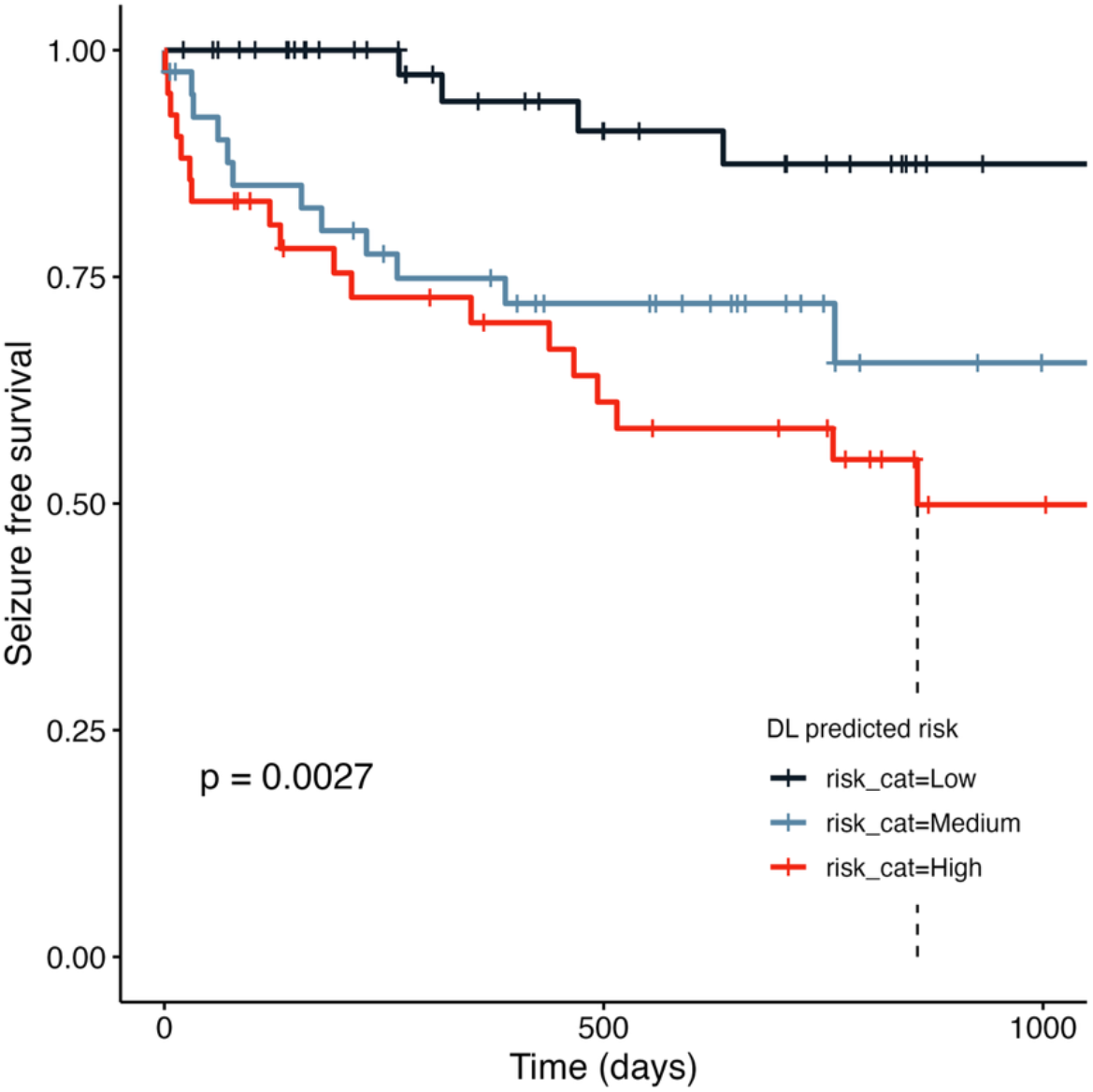
Seizure-free survival in the testing cohort, stratified the risk predicted by EEGSurvNet.

### Subgroup analyses

Stratified analyses reveal performance variations across subgroups (**Figure 6**). Young adults (<40 years) show better discrimination (two-year iAUROC: 0.73 [0.68-0.79]) then older subgroups (40-60 years: 0.64 [0.54-0.76]; > 60 year: 0.67 [0.58-0.82]). Regarding epilepsy type, the model performs better in focal epilepsy (0.66 [0.60-0.72]) then in generalized epilepsy (0.50 [0.39-0.77]), although this latter result is based on only 13 patients. Focal lesions and absence of abnormal slowing tend to improve performance.

**Figure 6:**
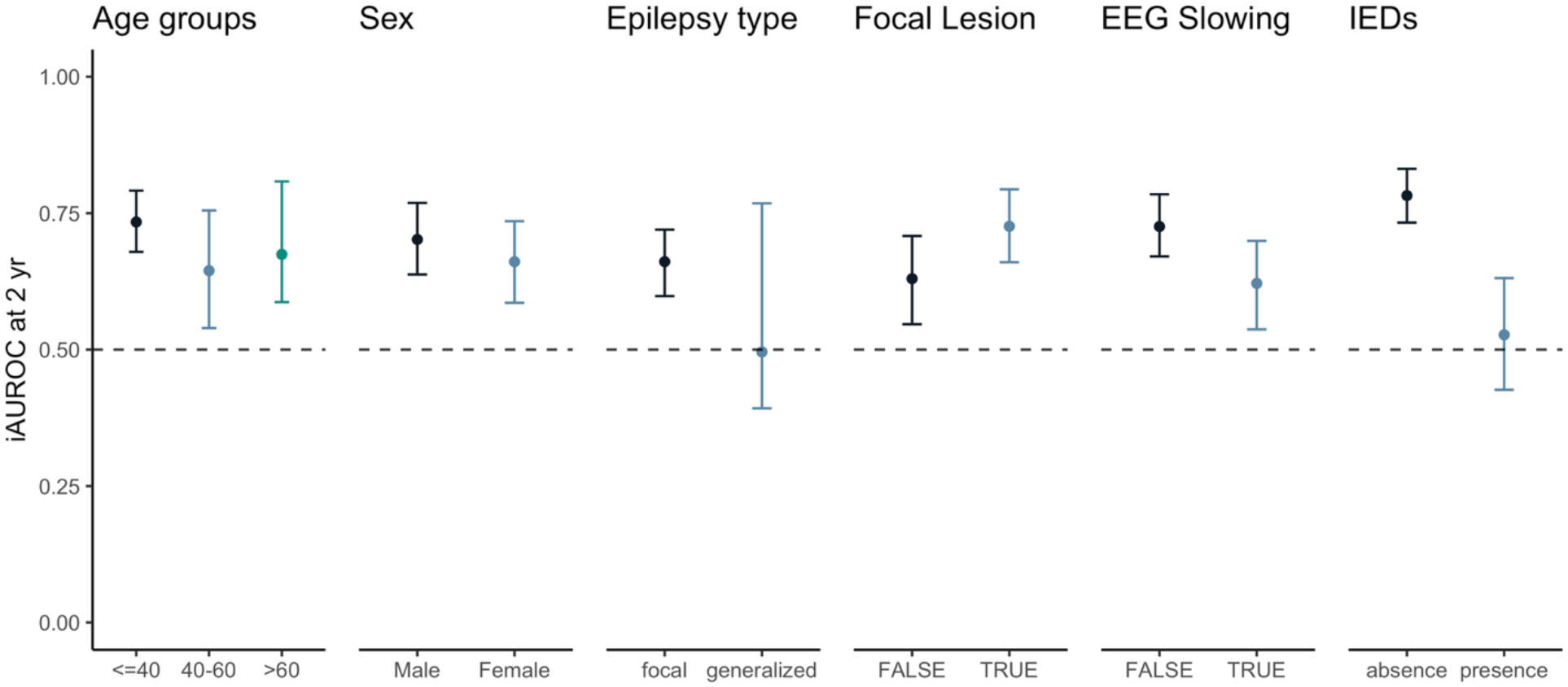
Analysis of predictive performance by subgroup. IED: Interictal epileptiform discharges.

The only statistically significant difference between subgroups concerns the presence of IEDs: the model performs better in their absence (iAUROC of 0.78 [0.73-0.83] vs.0.53 [0.43-0.63]).

### Model Interpretability

Shapley values were extracted for the 50 highest and 50 lowest predicted risk EEG segments (**Figure 7)**. The analysis demonstrates high importance of the 6–15 Hz frequency band. In terms of localization, channels in the temporal region show the highest importance, followed by occipital channels. A right-left asymmetry is also observed, with lower values in the right posterior temporal region.

**Figure 7:**
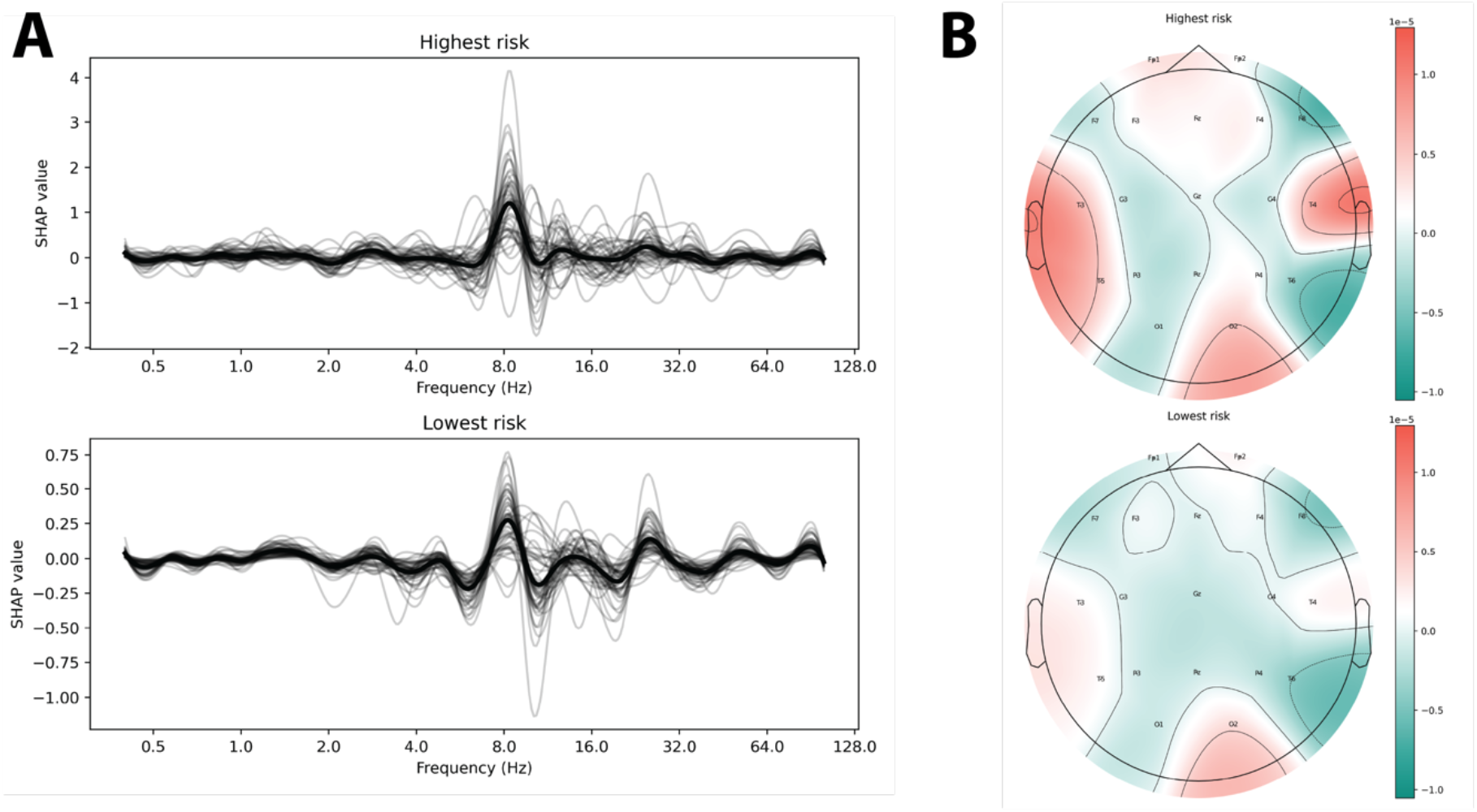
Shapley values for segments with highest and lowest predicted risk. A: Shapley values averaged by frequency band. The darker line corresponds to the average across 50 segments. B: Shapley values averaged by electrode.

### Ablation study

Results of the ablation study are presented in **Supplementary Table 3**. In all cases, performance is inferior to our model. The factor with the greatest impact is temporal resolution: when it is reduced by half, the two-year iAUROC drops from 0.69 to 0.54. The original DeepEpilepsy architecture, trained on raw signals rather than spectrograms, was not directly transferable to survival analysis (two-year iAUROC: 0.52).

## Discussion

This study presents the development and validation of a deep survival model, EEGSurvNet, to predict seizure risk from routine EEG. The model demonstrates superior performance compared to traditional predictors, both in terms of discrimination (two-year iAUROC = 0.69) and calibration (two-year iBS = 0.18). Performance is higher in the first months following EEG, reaching an AUROC of 0.80 at 2 months. The addition of clinical predictors marginally improves performance, suggesting that EEG captures essential prognostic information.

Survival analysis offers several advantages over traditional binary approaches. It explicitly models the temporal component of risk, accounts for variable follow-up durations, and allows simultaneous evaluation of effects at different intervals. Coupled with a deep model, it enables linking complex data like images, text, or time series with more granular clinical outcomes. Deep survival models have been used in oncology^24–26^ and intensive care^27^ to improve prognostic predictions and offer personalized therapeutic recommendations.

In epilepsy, traditional survival models have helped understand the disease’s natural history and identify prognostic factors.^42–44^ Validated models have been developed to predict seizure risk in various contexts, such as intracranial hemorrhage,^20^ ischemic stroke,^21,38^ cerebral venous thrombosis,^39^ head trauma,^40^ or after ASM withdrawal.^22^ However, despite this established use of survival models and the success of deep learning in other fields, no study has yet explored deep survival models for EEG analysis.

Existing prognostic models in epilepsy based on clinical variables such as IEDs, acute injury severity, or epilepsy duration, have been used in clinical practice.^45^ Their performance vary according to clinical context (C-indices: 0.67–0.89).^20–22,38–40^ EEGSurvNet’s performance is comparable to other prognostic models, particularly when combined with clinical data (C = 0.69). As with other predictive models in epilepsy, the real impact of these predictions on clinical decisions and patient outcomes remains to be demonstrated through prospective studies.^45^

EEGSurvNet performs better in young patients, those with focal epilepsy, EEGs without slowing, and EEGs without IEDs. Unfortunately, interpretation of this analysis is complicated by correlation between several of these variables in our sample: patients <40 years had predominantly focal epilepsy (92%), and patients with generalized epilepsy more frequently had IEDs (69% vs. 27% with focal epilepsy). Several hypotheses can still be proposed. Superior performance in young patients may reflect fewer confounding factors like polymedication or comorbidities. Better performance in focal epilepsy might reflect more marked signal abnormalities: 66% of patients had imaging lesions and 50% had abnormal EEG slowing, compared to only 8% and 15% respectively in generalized epilepsy, which could lead to more marked EEG signatures.

Notably, EEGSurvNet performed better in EEGs without epileptiform abnormalities (iAUROC: 0.78 vs 0.53). While partly explained by different epilepsy type (28% of EEGs in focal epilepsy showed IEDs vs. 69% in generalized epilepsy), it primarily indicates that the model uses signal characteristics distinct from classic epileptiform markers. The model’s output processing method, averaging predictions across the entire EEG, might reduce the weight of paroxysmal patterns like IEDs and favor more constant background rhythm abnormalities.

SHAP value analysis raises several hypotheses about alternative patterns detected by EEGSurvNet. Particular attention seems to be paid to temporal regions with left-right asymmetry, consistent with our cohort’s predominance of temporal lobe epilepsy (44% of cases, with predominant left lateralization). The importance of 6–15 Hz frequencies along a relatively high importance of occipital regions suggest reliance on posterior dominant rhythm, aligning with known alpha rhythm alterations in patients with epilepsy.^46–50^

This study has several limitations. First, all patients come from the same center. Despite clinical EEG standardization, certain factors may vary between centers, both in terms of recording and clinical practice. For example, our findings might poorly generalize to a general neurology practice with few refractory epilepsy cases, or a center where prolonged ambulatory EEG is more frequently used than routine EEG. Second, clinical follow-up differs between patients with and without epilepsy. Epilepsy patients tend to be followed over a longer timeframe, affecting the censoring probability in survival analysis. To mitigate this bias’s impact, we used inverse probability of censoring weights, but its real effect is difficult to quantify given that our data comes from real clinical practice. Third, our sample size remains modest for subgroup analyses, for which type II error cannot be excluded.

## Conclusion

In conclusion, this study demonstrates the performance of a deep survival model for predicting seizure risk up to two years after a routine EEG. Its ability to predict risk on EEGs without spikes reinforces the suspicion of neurophysiological biomarkers invisible to the naked eye, possibly in the 6-15 Hz frequencies.

This approach could greatly improve epilepsy diagnosis and monitoring, which are currently largely limited by the lack of quantifiable biomarkers of seizure risk. The next step will be to validate performance on samples from other centers. Subsequently, a prospective study of this model would allow evaluation of its real clinical utility in the diagnosis or monitoring of patients with epilepsy.

## Data Availability

The study source code will be available upon publication at: https://gitlab.com/chum-epilepsy/epi_surv. Anonymized data will be made available to qualified investigators upon reasonable request, conditional to the approval by our REB. The TRIPOD checklist is provided as Supplementary material.

## Acknowledgements

We would like to acknowledge the work of the CHUM EEG technologists for their contribution to the recording of the EEGs. We would also like to thank Manon Robert and Véronique Cloutier for their help regarding the access to the EEG data and the submission to the ERB.

## Author contributions

EL, DKN, FL, and EBA conceived and planned the experiments. EL, AQX, and MJ collected the data. EL, AQX, MJ, and EBA had direct access and verified the underlying data. EL performed the experiments. EL, DKN, FL, and EBA contributed to the interpretation of the results. EL wrote the first draft of the manuscript. All authors provided critical feedback and reviewed the manuscript.

