## Supplementary Material for "Development and Validation of a Deep Survival Model to Predict Time-to-Seizure from Routine EEG"

#### Supplementary Methods 1: EEGSurvNet Development and Specifications

EEGSurvNet is an improved version of our previous model, DeepEpilepsy, a Vision Transformer (ViT) that takes multi-channel EEG segments of 10 or 30s as input.^1^ The first improvement involves extending the duration of analyzed segments to 30 and 60 seconds. Beyond 60 seconds, we observed performance plateauing at the cost of very high computational resources. The second improvement applies a 60 Hz band-pass filter to eliminate power line noise contribution and potentially make the model robust to EEGs recorded in different countries. The third improvement concerns the input data format: EEGs are converted to spectrograms using Morlet wavelet transform (number of cycles: 7, number of frequency bands: 24). This improvement primarily enhances interpretability. Temporal resolution is decreased by a factor of 4 to limit input data size. The model's tokenizer was also modified to process these spectrograms, now using 2D convolutions. The final version has 27M parameters (**Supplementary Table 1**).

#### Supplementary Table 1: EEGSurvNet Specifications

| EEGSurvNet’s hyperparameter | Value |
| --- | --- |
| Patch size | 0.2s |
| Tokenizer: type | Convolution |
| Tokenizer: number of layers | 3 |
| Tokenizer: filter kernel size (frequency, time) | (3, 11) |
| Tokenizer: filter dimension | (256, 362, 512) |
| Transformer: hidden dimension | 512 |
| Transformer: number of layers | 8 |
| Transformer: number of heads | 8 |
| MLP: dimension | 1024 |
| Dropout | 0.2 |
| Params (M) | 27 |

### Supplementary Methods 2

We also tested a random reference model that predicts probabilities centered around baseline probabilities observed in the training set. Specifically, for each test set patient, the model generates a risk score drawn from a normal distribution ($\mu$ = mean training risk, $\sigma$ = 0.2). Model performance is compared to this reference via the integrated Brier score (iBS) calculated over 2 years, and the Brier Skill Score (BSS) at each period. BSS quantifies relative prediction improvement compared to the random model ^2^:

$$\text{BSS}(t)=1-\frac{\text{B}\text{S}_{\text{model}}(t)}{\text{B}\text{S}_{\text{ref}}(t)} ,$$

where $\text{B}\text{S}_{\text{model}}$ and $\text{B}\text{S}_{\text{ref}}$ are the Brier scores of the evaluated model and reference model (i.e., random model), respectively. BSS takes values between $]-\infty, 1.0]$, with positive values indicating performance superior to reference.

### Supplementary Methods 3

For model interpretation, we use Shapley values estimated via the gradient method, representing each model input value's contribution to its prediction.^47,48^ We trained the Shapley value estimator on 500 random segments from the training sample, then extracted values from the 50 segments with highest risk scores and 50 with lowest scores. Values are then averaged over time-frequency and spatial domains. The Python *shap* library was used for this analysis.^47^

Finally, we performed a systematic ablation study to evaluate the impact of four key model parameters: segment duration (30s vs. 60s), frequency resolution (16 vs. 32), temporal resolution (0.4s vs. 0.8s), and data augmentation (presence vs. absence). For each configuration, the model is retrained on the training set with fixed optimization parameters (learning rate, batch size, scheduling) and evaluated on the test sample. Additionally, the original DeepEpilepsy model^11^ was adapted for survival prediction by modifying its output layer to generate seven temporal risk values, then retrained on the training set.

#### Supplementary Table 2: Training Hyperparameters for EEGSurvNet

| Learning parameter | Value |
| --- | --- |
| Optimizer | AdamW |
| Maximal learning rate | 5.0 x10^-5^ |
| Weight decay | 0.05 |
| Momentum | β1, β2=0.9, 0.999 |
| Effective batch size | 512 |
| Number of GPUs | 2 |
| Epochs | 25 |
| Learning rate scheduler | *Cosine decay* |
| Warmup epochs | 1 |
| Warmup schedule | Linear |
| Augmentation probability | 0.9 |
| *Gradient clipping* | None |

#### Supplementary Table 3: Ablation Study for EEGSurvNet

| Model | Augmentation | Temporal resolution (s) | Frequency resolution | Segment duration (s) | iAUROC* |
| --- | --- | --- | --- | --- | --- |
| DeepEpilepsy | Oui | 0.05 | – | 30 | 0.52 |
| No_augment | Non | 0.2 | 32 | 60 | 0.58 |
| 16freq_8decim | Oui | 0.4 | 16 | 60 | 0.53 |
| 8decim | Oui | 0.4 | 32 | 60 | 0.54 |
| 16freq | Oui | 0.2 | 16 | 60 | 0.62 |
| 30s | Oui | 0.2 | 32 | 30 | 0.58 |
| EEGSurvNet | **Oui** | **0.2** | **32** | **60** | **0.69** |
| **Area under the ROC curve integrated over two years* | | | | | |
